## Supplemental Table 2 for "Improved detection of *SBDS* gene mutation by a new method of next-generation sequencing analysis based on the Chinese mutation spectrum"

Supplement Table 2 Fourfold table for the presence/absence of variant c.141C>T at PSV locus of *SBDS* gene and the corresponding variant chr7(GRCh37):g.72301284T>C at PSV locus of *SBDSP1* gene. All samples are from non-SDS group (N=20395). There is no statistically significant relationship between two variants (p>0.05).

|  | | chr7(GRCh37):g.72301284T>C  (*SBDSP1*) | | Total | Rate |
| --- | --- | --- | --- | --- | --- |
|  |  | Positive | Negative |  |  |
| NM_016038.2:c.141C>T  (*SBDS*) | Positive | 58 | 791 | 849 | 0.0416 |
|  | Negative | 950 | 18596 | 19546 | 0.9584 |
|  | Total | 1008 | 19387 | 20395 |  |
|  | Rate | 0.0494 | 0.9506 |  |  |
